# Interactive association between gut microbiota and thyroid cancer: a Mendelian randomization and systematic review

**DOI:** 10.1101/2023.03.27.23287832

**Authors:** Tianzhichao Hou, Qi Wang, Huajie Dai, Yanan Hou, Jie Zheng, Tiange Wang, Hong Lin, Shuangyuan Wang, Mian Li, Zhiyun Zhao, Yuhong Chen, Yu Xu, Jieli Lu, Ruixin Liu, Guang Ning, Weiqing Wang, Min Xu, Yufang Bi

**Affiliations:** Department of Endocrine and Metabolic Diseases, Shanghai Institute of Endocrine and Metabolic Diseases, Ruijin Hospital, Shanghai Jiao Tong University School of Medicine, Shanghai, China; Shanghai National Clinical Research Center for Metabolic Diseases, Key Laboratory for Endocrine and Metabolic Diseases of the National Health Commission of the PR China, Shanghai Key Laboratory for Endocrine Tumor, State Key Laboratory of Medical Genomics, Ruijin Hospital, Shanghai Jiao Tong University School of Medicine, Shanghai, China

**Keywords:** Thyroid Neoplasms, Gastrointestinal Microbiome, Causal Relationship, Mendelian Randomization Analysis

## Abstract

**Context:** The association between gut microbiota and thyroid cancer remains controversial.

**Objective:** We aimed to systematically investigate the interactive causal relationships between the abundance and metabolism pathways of gut microbiota, and thyroid cancer.

**Methods:** We leveraged the genome wide association studies for the abundance of 211 microbiota taxa from the MiBioGen study (N=18,340); 205 microbiota metabolism pathways from the Dutch Microbiome Project (N=7738); and thyroid cancer from the largest meta-analysis of Global Biobank Meta-analysis Initiative (N cases=6699 and N participants=1,620,354). We performed a bidirectional Mendelian randomization (MR) to investigate the causality from microbiota taxa, metabolism pathways to thyroid cancer, and vice versa. We did a systematic review of the previous observational studies and compared MR results with observational findings.

**Results:** Eight taxa and twelve metabolism pathways had causal effects on thyroid cancer, where *RuminococcaceaeUCG004* genus (*P* =0.001), *Streptococcaceae family* (*P* =0.016), *Olsenella* genus (*P* =0.029), ketogluconate metabolism pathway (*P* =0.003), pentose phosphate pathway (*P* =0.016), and L-arginine degradation II in AST pathway (*P* =0.0007) were supported by sensitivity analyses. Conversely, thyroid cancer had causal effects on three taxa and two metabolism pathways, where *Holdemanella* genus (*P* =0.015) was supported by sensitivity analyses. The *Proteobacteria* phylum, *Streptococcaceae* family, *Ruminococcus2* genus, and *Holdemanella* genus were significantly associated with thyroid cancer in both systematic review and MR, while other 121 significant taxa in observational results were not supported by MR.

**Discussions:** These findings implicated the potential role of host-microbiota crosstalk in thyroid cancer, while the discrepancy among observational studies called for further investigations.

## Introduction

The human body harbors many microbiota in the digestive tract, which plays a prominent role in gastrointestinal homeostasis, contributing to host conditions such as cardiometabolic diseases, autoimmune diseases, and neuro-psychiatric disorders. (1-3) Microbiome communities have been also suggested to influence the development, metastasis, and treatment response of multiple cancers, such as colorectal cancer, hepatic cancer, and gastric cancer.(4)

Thyroid carcinoma was also found to be associated with the alteration of gut microbiota abundance and microbiome dysbiosis.(5) Although past few decades have witnessed the incidence of thyroid cancer climb up to 6^th^ of all malignant tumors among females in the U.S. and 7^th^ in China, the underlying pathogenesis and risk factors of thyroid cancer remain unclear.(6,7) Previous studies indicated possible associations between gut microbiota and thyroid neoplasms. Studies from animal models demonstrated that thyroid homeostasis depended on the immune system and micronutrients, which were regulated by gut microbiota.(8) Sodium/iodide symporter in thyroid cells was also modulated by gut microbiome and contributed to papillary thyroid carcinoma through different mechanisms.(9) Besides, *Bacteroides fragilis* and *Bifidobacterium spp*. were associated with regulating the immune checkpoint, especially in thyroid cancer treatment.(10)

Nevertheless, results from observational studies varied largely, since a number of significant taxa could not well be replicated.(11-14) Due to the relatively insufficient recruited participants, these conclusions were prone to be influenced by various confounders and biases. Furthermore, limitations of cross-sectional studies made it difficult to determine whether thyroid cancer caused changes in microbiota composition, or gut microbiota was the initiator of the neoplasm.

We hereby aimed to comprehensively investigate the relationship between gut microbiota and thyroid cancer and discover potential underlined pathways based on the largest summary statistics so far in the bidirectional Mendelian randomization (MR). We also systematically reviewed previous observational studies on the same topic. Our results would illustrate a more explicit and complete correlation between host microorganism and thyroid cancer.

## Methods

### Study design

Firstly, we performed a large-scale MR to explore the causal effect of 211 human gut microbiota taxa and 205 microbiota-related metabolism pathways on thyroid cancer (Figure 1). Secondly, we performed MR from thyroid cancer to gut microbiota and metabolism pathways. Finally, we systematically reviewed all results of significant taxa change in patients with thyroid cancer from previous cross-sectional studies, and compared their results with the current study.

**Figure 1.**
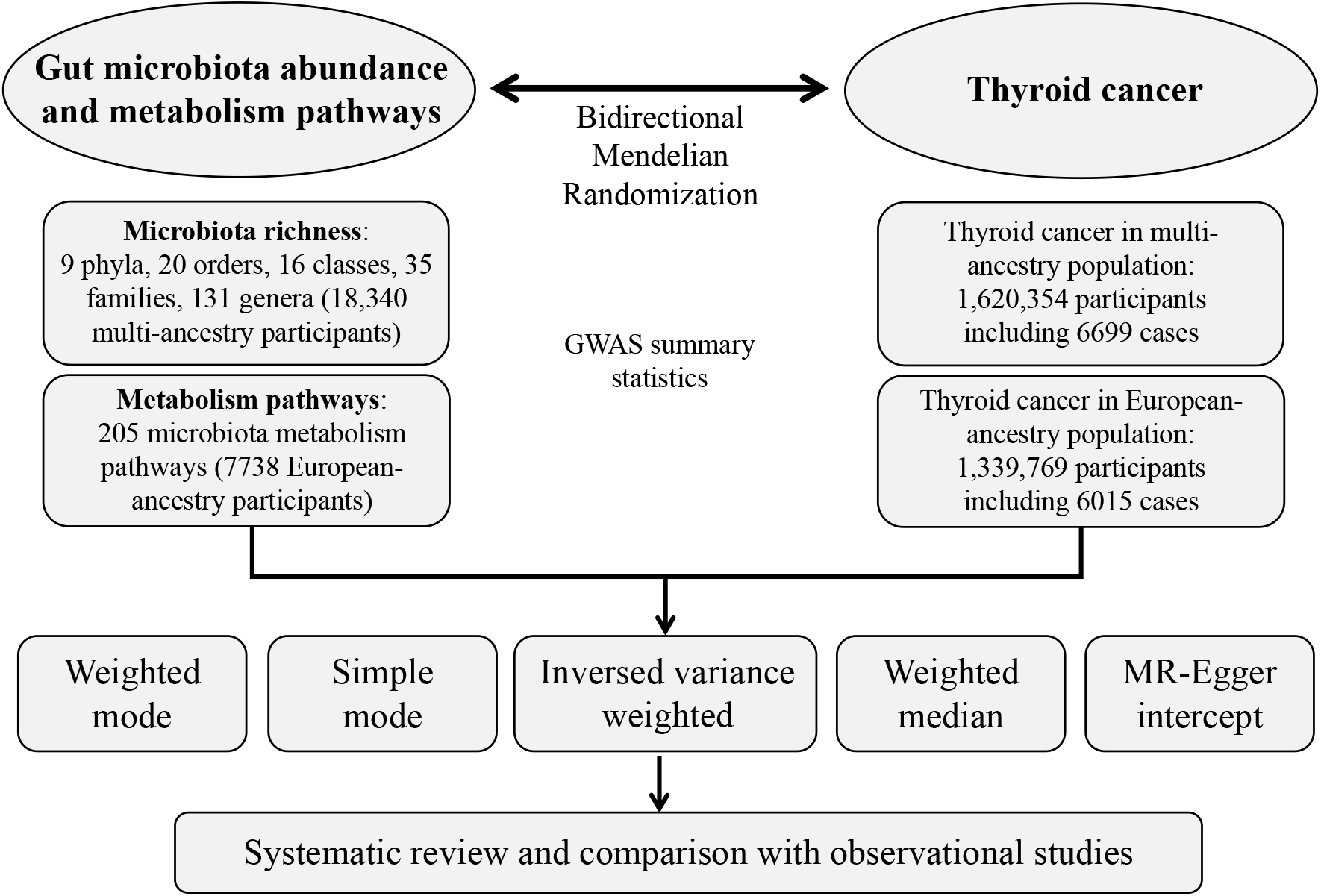
The flowchart and design of the bidirectional Mendelian randomization. This bidirectional MR was performed to investigate the causal effect of gut microbiota abundance, metabolism pathways on thyroid cancer, and vice versa. We then systematically reviewed results of significant taxa change in patients with thyroid cancer from previous cross-sectional studies, and compared their results with the current study.

This analysis was reported as the Strengthening the Reporting of Observational Studies in Epidemiology using Mendelian Randomization (STROBE-MR) guidelines (Table S1) and adopted several methods to follow the three fundamental assumptions of MR (15).

### Exposure and outcome selection

The data source of phenotypes was shown in Table 1. The datasets related to microbiota and metabolism pathways were extracted from two studies. Microbiota was from the largest genome-wide association study (GWAS) of host genetic variation of gut microbiota abundance in 18,340 multiple-ancestries participants (85% European-ancestry) from 24 independent cohorts in the MiBioGen consortium.(16) Based on the 16S ribosomal RNA gene sequencing, it offered GWAS summary statistics of 211 taxa: 9 phyla, 16 classes, 20 orders, 35 families, and 131 genera.

**Table 1.** Summary data sources of the study.

| Phenotype | Detail info. | Sample size | Ancestry | Available year | Dataset source | Reference |
| --- | --- | --- | --- | --- | --- | --- |
| Microbiota taxonomy | 211 gut microbiota taxa | 18 340 | Multiple (85% Eur) | 2021 | MiBioGen | Kurilshikov et al.(16) |
| Microbiota metabolism pathway | 205 metabolism pathways | 7738 | Eur | 2022 | DMP | Lopera et al.(17) |
| Host thyroid cancer (ThC) | ThC from 13 biobanks | 1 620 354 (6699 cases) | Multiple (83% Eur) | 2021 | GBMI | Zhou et al.(18) |
| Host thyroid cancer (ThC) | ThC from 11 biobanks | 1 339 769 (6015 cases) | Eur | 2021 | GBMI | Zhou et al.(18) |

Abbreviation: DMP: Dutch Microbiome Project, GBMI: Global Biobank Meta-analysis Initiative

Bacterial pathways were from a GWAS representing microbial function using shotgun metagenomic sequencing in 7738 European-ancestry participants of the Dutch Microbiome Project.(17) We used 205 bacteria metabolism pathways from Dutch Microbiome Project in the subsequent analysis.

The thyroid cancer was from the GWAS summary statistics released by Global Biobank Meta-analysis Initiative (GBMI).(18) It is the largest meta-analysis collaborative network of 19 biobanks from four continents representing over 2.1 million consented individuals with genetic data linked to electronic health records. The thyroid cancer was defined as either “ICD10 codes include all subcategories of the code” or “self-reported thyroid cancer”. We used the thyroid cancer GWAS summary statistics of both multi-ancestry data (N = 1,620,354, 83% European-ancestry, Case =6699) and European-ancestry data (N = 1,339,769, Case =6015). In the bidirectional MR analysis, the causality between thyroid cancer and microbiota abundance was performed based on multi-ancestry data from the MiBioGen consortium; the causality between thyroid cancer and metabolism pathways abundance was performed based on European-ancestry data from the Dutch Microbiome Project.

### Statistics analysis

The exposure of each taxon and pathway of microbiota used the following process to select IVs. Consistent with previous studies, (19,20) single nucleotide polymorphisms (SNPs) with genome-wide significance *P* <1×10^− 5^ and effect allele frequency (EAF) >0.01 were included. We also calculated the *F*-statistics to avoid weak instrument bias. All these genetic variants were clumped to a linkage disequilibrium (LD) threshold of r^2^ <0.001 for multi-ancestry microbiota taxa and European-ancestry microbiota pathway using the 1000 Genomes European reference panel as we previously described.(21-24) Furthermore, we used proxies of SNPs as substitutes with r^2^ ≥0.8 if the SNPs could not be found in the thyroid cancer GWAS summary data.(25) To avoid any bias due to IVs pleiotropy, we performed a Phenome-wide association study of all used SNPs in the IEU Open GWAS Project, and removed any SNPs that significantly related to thyroid cancer. Finally, the remaining genetic variants were used as IVs to model the effect of specific taxon and pathway of gut microbiota on thyroid cancer.

SNPs of thyroid cancer with genome-wide significance *P* <5×10^− 8^ (for both multi-ancestry and European-ancestry data) and EAF >0.01 were included. Similar to exposure of gut microbiota, all SNPs were clumped to a LD threshold of r^2^ <0.001 using the 1000 Genomes European reference panel.(26) We obtained the MR estimates for the mutually causal effect of using the inverse-variance weighted (IVW) method. The estimate was provided as odds ratio (OR) with 95% confidence interval [95% CI]. MR results with *P* <0.05 were considered nominally significant. We also used Benjamini-Hochberg method of false discovery rate (FDR) correction for multiple testing. Heterogeneity of effects was assessed by scatter plots and Cochran’s Q test for the SNP exposure and outcome associations. We also performed sensitivity analyses using the Weighted median, MR-Egger, and mode-based methods to validate the results from the IVW method.(27,28) Egger intercept was used to assess the horizontal pleiotropy and any results whose *P* value of Egger intercept <0.05 were excluded.

We then estimated the heritability of 211 microbiota taxa using single-variate linkage disequilibrium score regression (LDSC) and genetic correlation between gut microbiota and thyroid cancer via bivariate LDSC for all high-quality genetic variants (i.e., INFO score > 0.9) from each GWAS. All MR analyses were based on the TwoSampleMR package in R software, version 4.1.2.(25)

### Systematic literature review and comparison

We carried out a systematic review to of observational studies to compare with our MR results in accordance with the Preferred Reporting Items for Systematic Reviews and Meta-Analyses (PRISMA) statement. The inclusion criteria were observational studies assessing the gut microbiota taxa change associated with thyroid cancer, with available linear discriminant analysis (LDA) score. The LDA effect size is an order of magnitude difference in abundance, which reveals the statistically significant taxa between groups. Studies assessing taxa change in only thyroid nodule, the control groups were not healthy adults, conference abstracts, grey literature, and unpublished studies were excluded.

We search for relevant publications based on “Thyroid Neoplasms”[MeSH] AND “Gastrointestinal Microbiome”[MeSH] in PubMed and ‘thyroid cancer’ AND ‘intestine flora’ in Embase. The search was performed in January 2023. All studies from the databases were exported. Duplicates were removed, before titles, abstracts, and figures were screened by two authors (T.H. and Q.W.), for potentially eligible studies. Full texts were subsequently assessed and discrepancies during the study selection process were resolved via discussions with a third investigator (H.D.). We extracted the LDA score, which represented the abundance of taxon in the host, of significant taxa in our MR results from the main text or supplements. We compared our MR results with LDA score showed in the observational studies.

## Results

### Exposure SNPs selection

For 211 taxa in the MiBioGen consortium, the remaining genetic variants for each taxon exposure ranged from 4 to 26 SNPs (median 13 SNPs, F-statistics = 21) (Table S2). For 205 pathways in Dutch Microbiome Project, the remaining genetic variants ranged from 1 to 20 SNPs (median 11 SNPs, F-statistics = 25) (Table S3). The exposure finally remained 23 SNPs for multi-ancestry thyroid cancer and 19 SNPs for European-ancestry thyroid cancer (Table S4).

### MR estimates for causal effect of gut microbiota and pathways on thyroid cancer

Based on the IVW method, MR results supported causal effects of gut microbiota composition and metabolism pathways on thyroid cancer. We identified that eight taxa and 12 metabolism pathways were causally correlated with thyroid cancer risk (Figure 2, Table S5-S6).

**Figure 2.**
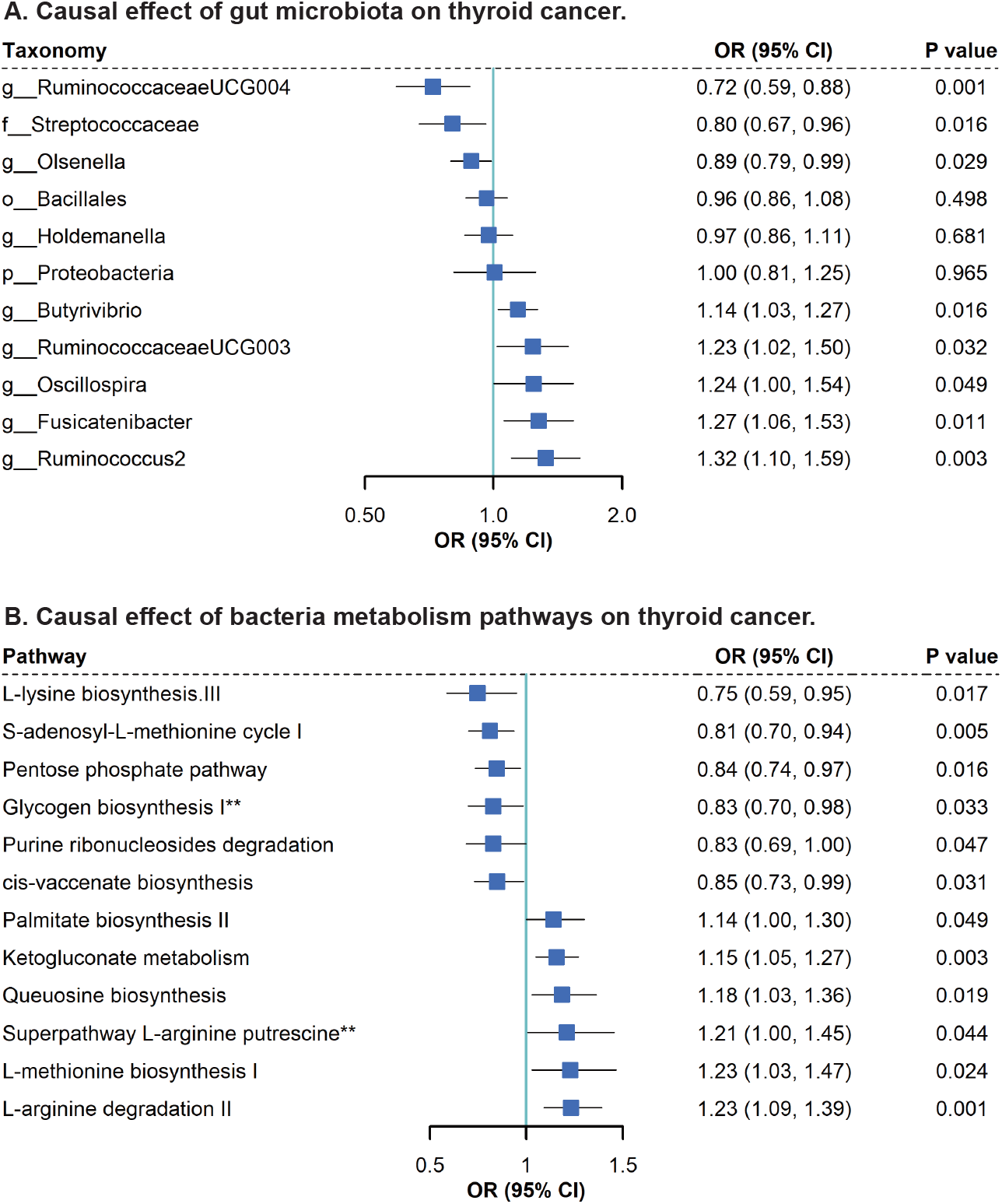
Causal effect of gut microbiota taxa and metabolism pathways on thyroid cancer. **A**. Causal effect of gut microbiota taxa on thyroid cancer, including 8 taxa with significant effect on thyroid cancer and 3 non-significant taxa. **B**. Causal effect of 12 gut microbiota metabolism pathways on thyroid cancer. **The whole names are glycogen biosynthesis I from ADP-D-Glucose and superpathway of L-arginine, putrescine, and 4-aminobutanoate degradation. Abbreviation: the prefix “g” represents for genus, “f” for family, “o” for order, “c” for class, and “p”: phylum.

From microbiota composition to thyroid cancer, we found that per unit increased abundance of *RuminococcaceaeUCG004* genus (OR: 0.72, 95% CI: 0.59-0.88, *P* =0.001), *Ruminococcus2* genus (OR: 1.32, 95% CI: 1.10-1.59, *P* =0.003), *Fusicatenibacter* genus (OR: 1.27, 95% CI: 1.06-1.53, *P* =0.011), *Butyrivibrio* genus (OR: 1.14, 95% CI: 1.03-1.27, *P* =0.016), *Olsenella* genus (OR: 0.89, 95% CI: 0.79-0.99, P =0.029), *RuminococcaceaeUCG003* genus (OR: 1.23, 95% CI: 1.02-1.50, *P* =0.032), *Oscillospira* genus (OR: 1.24, 95% CI: 1.00-1.54, *P* =0.049), and *Streptococcaceae* family (OR: 0.80, 95% CI: 0.67-0.96, *P* =0.016) were causally associated with thyroid cancer risk. All 2818 SNPs of bacteria were related to 16,603 phenotypes under a threshold of 1×10^-5^, where none of SNPs were related to the thyroid cancer in the PhenoWAS study. Three non-significant taxa, *Subdoligranulum genus, Romboutsia genus, Adlercreutzia genus*, with Egger intercept *P* <0.05 had been excluded, which indicated little pleiotropy in the rest results.

From microbiota metabolism pathways to thyroid cancer, we found that per unit increased abundance of 12 pathways, including L-arginine degradation II in AST pathway (OR: 1.23, 95% CI: 1.09-1.30, *P* <0.001), ketogluconate metabolism (OR: 1.15, 95% CI: 1.05-1.27, *P* =0.003), S-adenosyl-L-methionine cycle I (OR: 0.81, 95% CI: 0.70-0.94, *P* =0.005), pentose phosphate pathway (OR: 0.84, 95% CI: 0.74-0.97, *P* =0.016), L-lysine biosynthesis III (OR: 0.75, 95% CI: 0.59-0.95, *P* =0.017), Queuosine biosynthesis (OR: 1.18, 95% CI: 1.03-1.36, *P* =0.019), L-methionine biosynthesis I (OR: 1.23, 95% CI: 1.03-1.47, *P* =0.024), cis-vaccenate biosynthesis (OR: 0.85, 95% CI: 0.73-0.99, *P* =0.031), glycogen biosynthesis I from ADP -D-Glucose (OR: 0.83, 95% CI: 0.70-0.98, *P* =0.033), superpathway of L-arginine putrescine and 4-aminobutanoate degradation (Wald Ratio OR: 1.21, 95% CI: 1.00-1.45, *P* =0.044), purine ribonucleosides degradation(OR: 0.83, 95% CI: 0.69-0.99, *P* =0.047), and palmitate biosynthesis II (OR: 1.14, 95% CI: 1.00-1.30, *P* =0.049) had a causal effect on thyroid cancer. Five metabolism pathways with intercept *P* <0.05 were excluded.

Although IVW results indicated none of the taxon or pathway was significant after FDR correction, weighted median method still supported the significant results of *RuminococcaceaeUCG004* genus, *Olsenella* genus, and *Streptococcaceae* family, ketogluconate metabolism, pentose phosphate pathway, and L-arginine degradation II in AST pathway.

### MR estimates for causal effect of thyroid cancer on gut microbiota and pathways

Based on the IVW method, MR results supported causal effects of thyroid cancer on three taxa and two bacterial metabolism pathways (Figure 3, Table S7-S8).

**Figure 3.**
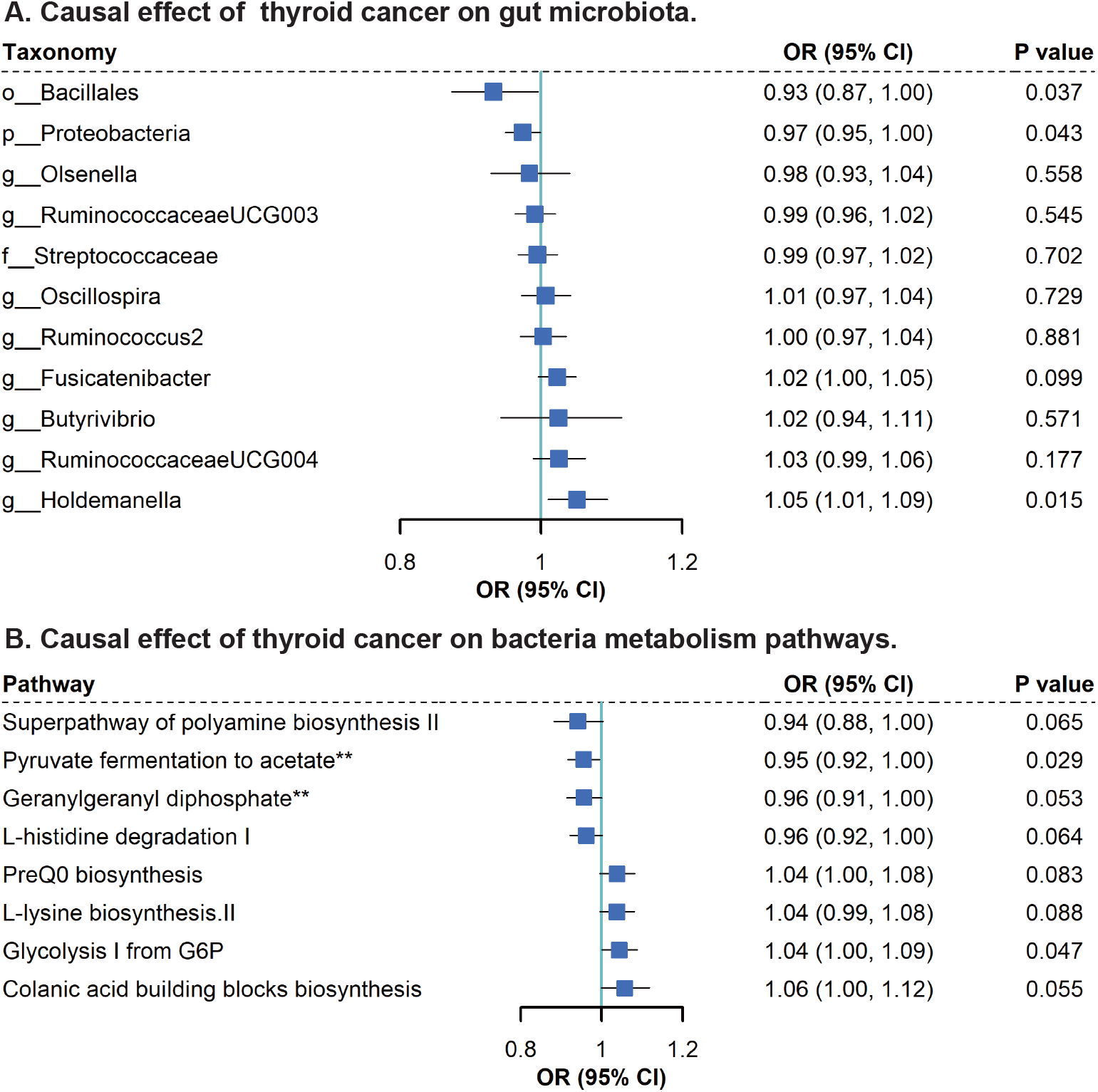
Causal effect of thyroid cancer on gut microbiota taxa and metabolism pathways. **A**. Causal effect of thyroid cancer on gut microbiota taxa, including 3 significant taxa and 3 non-significant taxa. **B**. Causal effect of thyroid cancer on 2 significant metabolism pathways and 6 non-significant pathways. **The whole names are superpathway of polyamine biosynthesis II and superpathway of geranylgeranyl diphosphate biosynthesis II via MEP. Abbreviation: the prefix “g” represents for genus, “f” for family, “o” for order, “c” for class, and “p”: phylum.

For the thyroid cancer to gut microbiota, thyroid cancer increased abundance of *Holdemanella* genus (OR: 1.05, 95% CI: 1.01-1.09, *P* =0.015) and decreased abundance of *Bacillales* order (OR: 0.93, 95% CI: 0.87-1.00, *P* =0.037), and *Proteobacteria* phylum (OR: 0.97, 95% CI: 0.95-1.00, *P* =0.043) in the multi-ancestry analysis. For the thyroid cancer to microbiota-related pathways, we found thyroid cancer had causal effects on *pyruvate fermentation to acetate and lactate II* (OR: 0.95, 95% CI: 0.92-1.00, *P* =0.029), and *glycolysis I from glucose-6-phosphate* (OR: 1.04, 95% CI: 1.00-1.09, *P* =0.047). Taxa and pathways with Egger intercept *P* <0.05 were excluded, and all results mentioned above had little pleiotropy.

Although IVW results showed none of the taxon or pathway was significant after FDR correction, the MR-Egger and weighted median methods both supported significance of *Holdemanella* genus. The comparison of the effect of gut microbiota on thyroid cancer with thyroid cancer on gut microbiota was shown in Figure 4.

**Figure 4.**
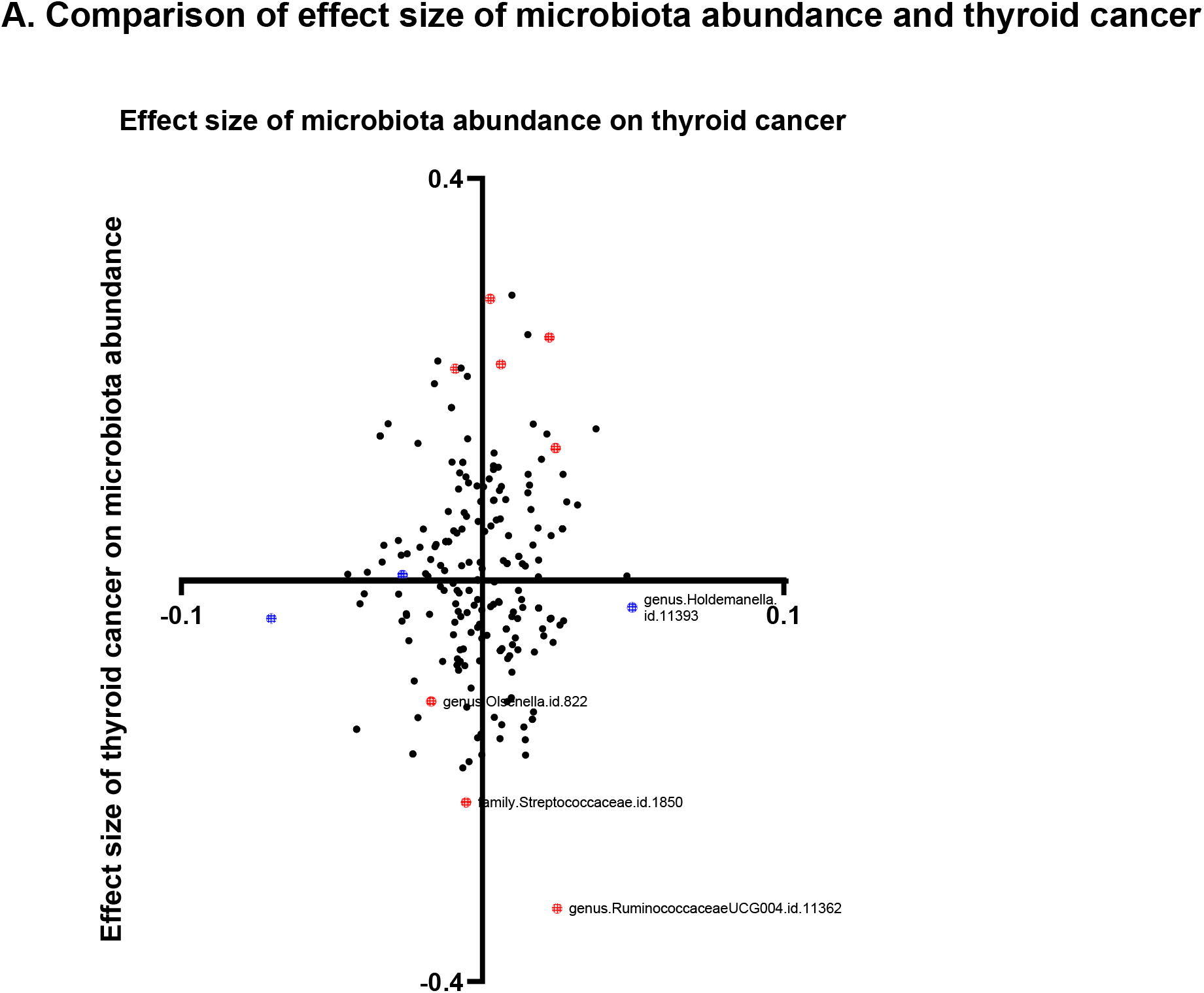
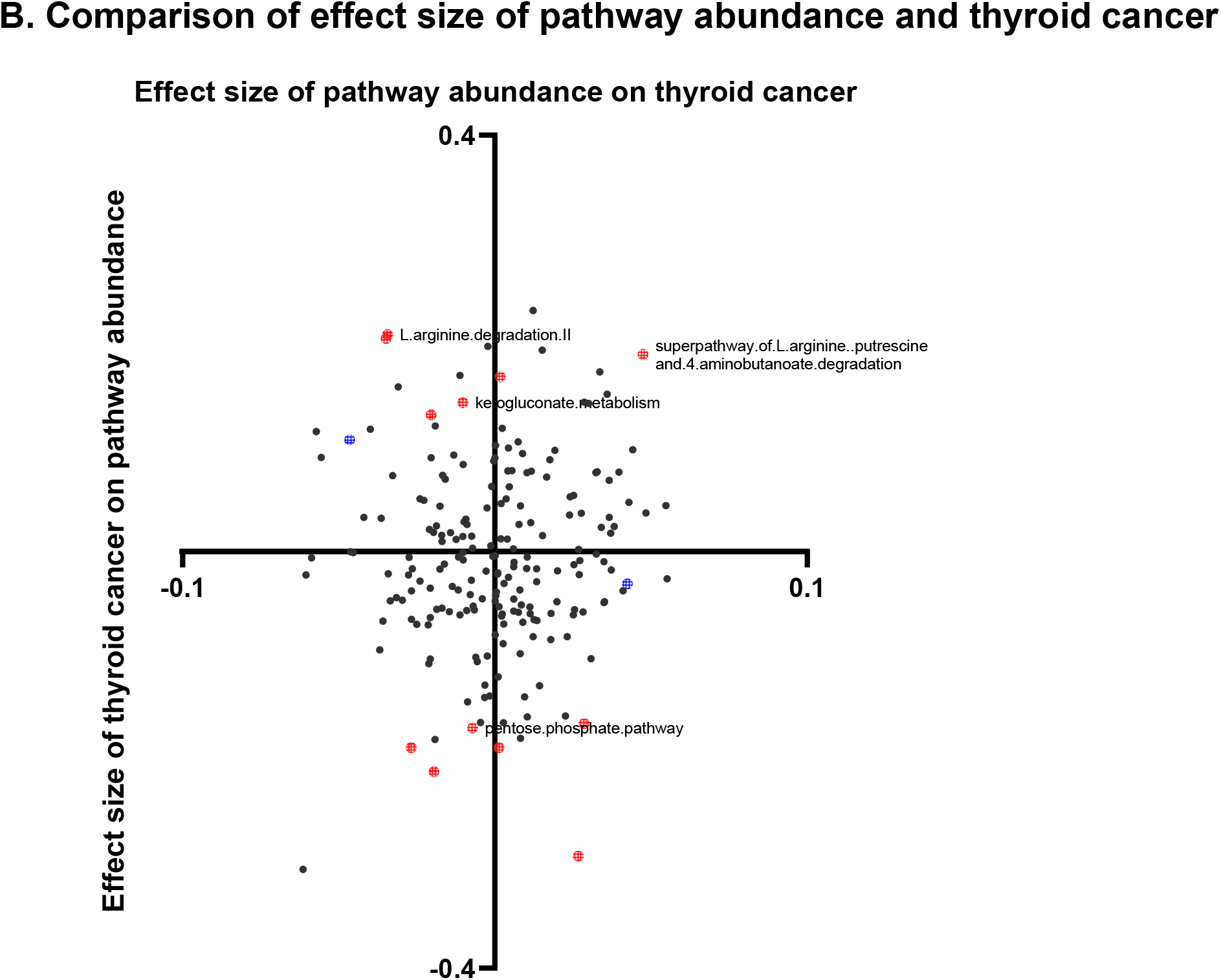
Comparison of the effect size of bidirectional MR. **A**. Comparison of the effect size of thyroid cancer on microbiota abundance with the effect size of microbiota abundance on thyroid cancer. **B**. Comparison of the effect size of thyroid cancer on pathway abundance with the effect size of pathway abundance on thyroid cancer. Red points highlighted significant results for microbiota/pathway on thyroid cancer and blue points was for thyroid cancer on microbiota/pathway.

From the single-variate LDSC, the heritability of each taxon ranged from 0.6% (*Eubacteriumrectalegroup* genus) to 20.6% (*Sellimonas* genus). However, no significant genetic correlation was detected in bivariate LDSC (Table S10).

### Systematic review and comparison with observational studies

Our research strategy identified totally 55 studies from both PubMed (10 records) and Embase database (45 records) (Figure 5). We excluded 8 duplicated records and 27 non-article studies labeled in Embase. After exclusion in title & abstract screening, ten studies remained for full text screening and finally, we included 4 studies with available LDA score of gut microbiota abundance in thyroid cancer patients compared with healthy participants. (11-14) There were minimal discrepancies between authors in the duplication of the search strategy.

**Figure 5.**
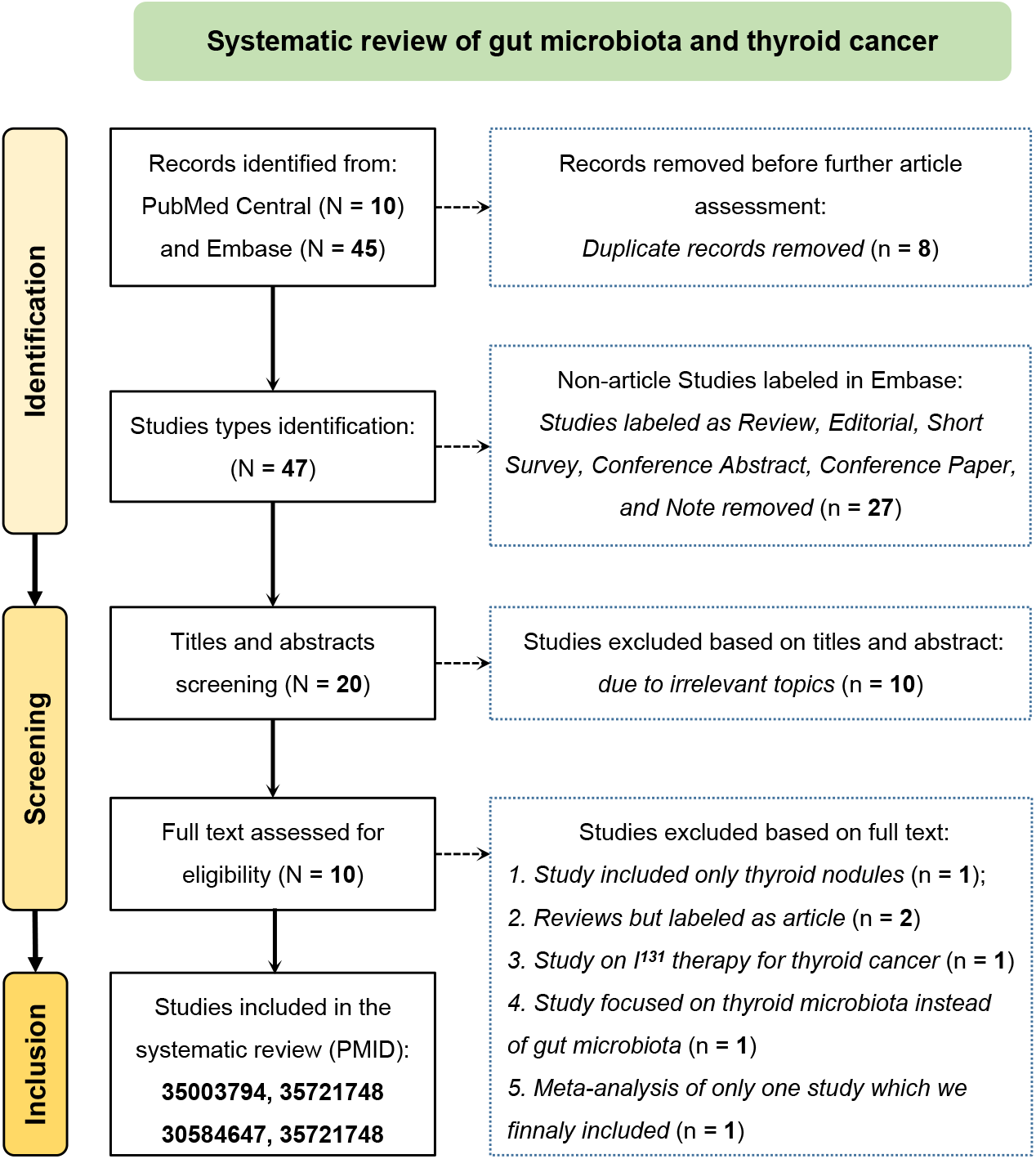
Flow diagram of screening process of studies in PubMed and Embase included in systematic review. A total of 55 records focusing on gut microbiota and thyroid cancer were found in PubMed and Embase. After the exclusion and inclusion criteria, four studies remained in the final systematic review.

All these 4 studies were cross-sectional studies performed in the Chinese population based on 16S rRNA sequencing. A total of 125 taxa with available LDA score were reported. Taxa with LDA score greater than the set value, the default cutoff values were 2 [log 10] for Zhang et al.,(13) 3 for Feng et al. & Lu et al., (12,14) and 3.5 for Yu et al.(11) We searched microbiota with significant MR results in the observational studies and compared MR effect size with significant LDA score reported. Among the 11 taxa with significant MR results, 4 were reported with significant LDA scores in observational studies, including *Proteobacteria* phylum, *Streptococcaceae* family, *Ruminococcus2* genus, and *Holdemanella* genus (Figure 6).

**Figure 6.**
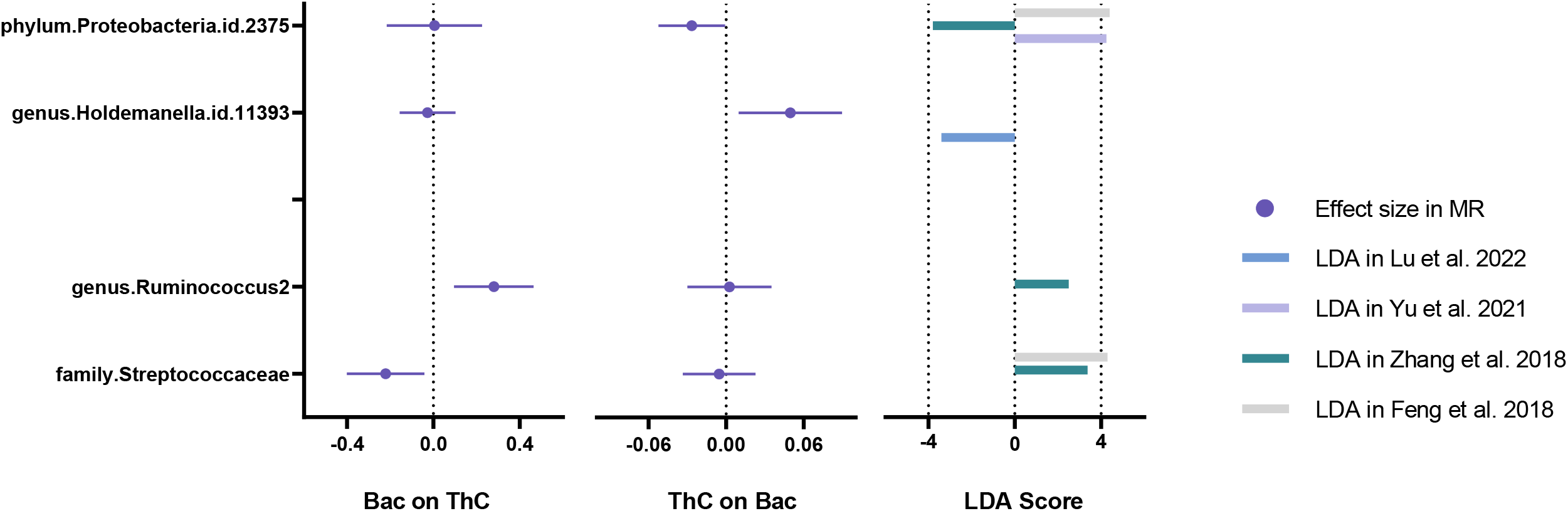
The forest plot of bidirectional MR results compared with observational results in the systematic review. Comparison of four taxa of significant MR results with four observational LDA score. The abundance of *Proteobacteria* phylum and *Holdemanella* genus (upper two taxa) were significantly affect by the presence of thyroid cancer; the abundance of *Ruminococcus2* genus and *Streptococcaceae* family (lower two taxa) significantly affected thyroid cancer. Abbreviation: Bac: Bacteria (gut microbiota abundance), ThC: thyroid cancer;

MR showed a positive effect of *Ruminococcus2* genus on thyroid cancer, which was consistent with the observational findings.(13)Thyroid cancer had a negative effect on *Proteobacteria* phylum, which was supported by Zhang et al.,(13) while other 2 studies showed an increased abundance of *Proteobacteria* phylum in thyroid cancer patients. (11,12) *Streptococcaceae* family had a negative effect on thyroid cancer in MR, while thyroid cancer patients had an elevated abundance of the taxon in observational results.(12,13) Although thyroid cancer had a negative effect on *Holdemanella* genus, Lu et al. found an increased abundance of *Holdemanella* genus in thyroid cancer patients. For other 121 taxa that significantly reported to be associated with thyroid cancer, we did not observe evidence of significant causal relationship in MR.

## Discussion

Based on the largest GWAS of thyroid cancer and gut microbiota, the present bidirectional MR assessed the potential genetic causality between the abundance and metabolic pathways of gut microbiota and thyroid cancer. Eight taxa and twelve pathways had causal effects on thyroid cancer; and thyroid cancer had causal effects on three taxa and two pathways. The systematic review showed 125 results of taxa change in observational studies, where *Proteobacteria* phylum, *Streptococcaceae* family, *Ruminococcus2* genus, and *Holdemanella* genus could be found with significance in MR (other 121 results were not significant). Our findings implicated the potential role of host-microbiota crosstalk related to thyroid cancer; however, given the large discrepancy among all observational studies, and the difference between MR and systematic review, the causality between the reported taxa and thyroid cancer were still warranted further validation.

Recently, the associations between gut microbiota and thyroid disorders have been increasingly investigated, while still poorly understood.(8) Four cross-sectional studies were found and finally included in the systematic review.(11-14) However, the interpretation of these studies was limited to their inadequate participants and the shortness of study design. Cross-sectional studies could reveal the correlation but not directional causality. The change of bacteria composition in patients with thyroid cancer could be attributed to pathogen causing neoplasm, caused by neoplasm, or only accompanied with neoplasm presence. Therefore, since non-genetic and genetic factors each account for similar variation in gut microbiota (approximately 10%),(29) directional causality inferred from genetic evidence, for instance MR, was beneficial to further validate and clarify observational results.

Among elven significant taxa in MR, the change of *Proteobacteria* phylum and *Streptococcaceae* family could be found in more than one observational study. Bidirectional MR illustrated that the abundance of *Proteobacteria* phylum, considered a signature of microbial dysbiosis, was lower in thyroid cancer patients, which was consistent with what was found in a cohort of 74 patients with thyroid neoplasm.(13) Other three studies investigated the relationship between dysbiosis of gut microbiota and thyroid cancer in Chinese participants and highlighted an increased abundance of *Proteobacteria* phylum in thyroid cancer patients.(11,12,14) The controversial results indicated that *Proteobacteria* phylum was a complicated and variable composition contributing to dysbiosis in gut microbiota. Additionally, *Streptococcaceae* family might decrease the risk of thyroid cancer in MR, while cross-sectional studies showed an increased abundance in thyroid cancer patients. The discrepancies between single-center cross-sectional studies and MR could be attributed to the small sample size (<100), non-chronologic non-causal investigations, or various populations (European vs. Chinese). Furthermore, MR showed the causality of a lifelong exposure on the outcome, while cross-sectional studies focused on a specific age when the research was performed. Given that age represents the strongest factor that can account for variation in gut microbiota,(30) results of lifelong exposure in MR would be different from results in cross-sectional time in observational studies.

However, most of taxa with significant change of abundance in the four studies (121 from 125 results) could hardly be replicated in the bidirectional MR. This suggested that the previously reported associations between gut microbiota and thyroid cancer may not always underly causal relations. Instead, the abundance change of gut microbiota might be an accompaniment with host diseases status.

Gut microorganisms could also influence tumorigenesis via multiple intra-bacterial metabolism pathways through the gut-thyroid axis.(8) These pathways included secretion of virulence factors, physical binding induced signaling, and immune cell recruitment.(4) Queuosine, mostly derived from *E. coli*, was able to alter t-RNA base and promote the proliferation of malignant cells of multiple cancer types by inducing unfolded protein response.(31) In addition, bacterial amino acid metabolism was related to host amino acid metabolism and thyroid cancer progress.(32) Bacterial

L-arginine degradation could weaken the host immune T cell response, contributing to cancer development, which was a feasible target for cancer therapy.(33-35) Besides, some bacterial pathways might ameliorate host immune response and reduce cancer risk. MR indicated S-adenosyl-L-methionine (SAM) cycle I had a significant causal effect on decreasing thyroid cancer risk, which was consistent with experimental studies in other types of cancers.(36) SAM induced a marked cell cycle arrest at the G2/M phase and downregulated pro-metastatic genes in squamous cancer cells.

Therefore, the association of microbiota metabolism with thyroid cancer required further investigated. The strength of this study took advantage of the large sample size of both thyroid cancer and microbiota with adequate power and no sample-overlap. Especially, the GBMI for thyroid cancer was the largest meta-analysis of 19 global biobanks.

Besides, several sensitivity analyses and the systematic review showed the replicated results with the MR, which provided consistent evidence for the relation between gut microbiota and thyroid cancer. The limitations should also be acknowledged. Firstly, the use of a significant threshold of 1×10^-5^ may be subject to challenge. However, this threshold was based on the original GWAS of MiBioGen, (16) and subsequent MR studies, (19,20) which was used to increase the number of SNPs available for sensitivity analyses. In addition, the *F*-statistics of the IVs is sufficient (all ≥20), and it could largely limit the potential weak IV bias. Secondly, the 16S rRNA gene sequencing was used in the determination of the gut microbiota taxa and metabolic pathways in both MR and the systematic review literatures, which allowed resolution only from the genus to the phylum level. It obscured identification the potentially significant associations of individual species with thyroid cancer. Thirdly, given the diverse and less precise definition of thyroid cancer across studies included in the GBMI, it was challenging to conduct MR on subtypes such as follicular cancer or undifferentiated cancer. Finally, the microbiota composition was influenced by different ancestries, and the genetics of gut microbiota is highly heterogeneric. The interpretation of MR results to other ancestries should be with carefulness.

## Conclusion

This is the first study comprehensively assessing the relationship between gut microbiota and thyroid cancer. Eight taxa and twelve metabolism pathways had causal effects on thyroid cancer; and thyroid cancer had causal effects on three taxa and two metabolism pathways. Four taxa in systematic review were found with significant MR results, including *Proteobacteria* phylum, *Streptococcaceae* family, *Ruminococcus2* genus, and *Holdemanella* genus. Our results suggested the potential role of gut microorganisms in host thyroid cancer and implicated the importance of modulating host-microbe balance in the cancer prevention and treatment. However, due to the discrepancy among observational studies and MR, the association of previously reported taxa with thyroid cancer was suspected and thus, larger prospective studies were still warranted.

## Data Availability

All data sources are available at Table 1

## Acknowledgements

We deliver our thanks to Miss Mingqing Yuan from Zhejiang University for her figure making and Dr. Yunye He from Tokyo University for method guiding. We want to acknowledge all participants in the GWAS study.

## Availability of data and materials

Data availability can be found in Table 1.

## Authors’ contributions

M.X., T.H., Y.B., and W.W. conceived and designed the study; T.H., Q.W., H.D., and Y.H. conducted the analysis and finished the paper writing; J.Z., T.W., M.L., H.L., S.W., Z.Z., R.L., and Y.X. offered guidance and methods for the analysis and data selection; Y.C., J.L., W.W., G.N., and Y.B. reviewed the article and offered clinical advices. M.X., Y.B., and W.W. take responsibility for the contents of the article. All authors read and approved the final version of the manuscript.

## Ethics approval and consent to participate

Ethics consent has been approved by original studies.

